# Risk of adverse outcomes with COVID-19 in the Republic of Ireland

**DOI:** 10.1101/2020.12.09.20246363

**Authors:** M Roe, P Wall, P Mallon, M Horgan

## Abstract

**Aims:** To compare the risk of adverse outcomes (i.e. hospital/intensive care admission, death) in population sub-groups during two periods of the COVID-19 pandemic in the Republic of Ireland.

**Methods:** We analysed routinely-collected, publicly-available data on 67,900 people with laboratory confirmed COVID-19 infection between 29^th^ Feb to 14^th^ Nov 2020. This period encompassed two waves of infection and two corresponding national lockdowns. For two observational periods covering each wave (W1, W2), each ending 17-19 days before implementation of high-level national restrictions, we segmented the population based on age and underlying clinical conditions.

**Results:** The prevalence of laboratory confirmed COVID-19 was 1.4%. The risk of admission to hospital, admission to intensive care, and death was 7.2%, 0.9%, and 2.5%, respectively. Compared to younger confirmed cases, those aged ≥65 y had increased risk of hospital admission (RR 5.61), ICU admission (RR 3.56), and death (RR 60.8). W2 was associated with more cases and fewer adverse events than W1. The risk of all adverse outcomes was reduced in W2 than in W1.

**Conclusions:** Ongoing responses should consider the variation in risk of adverse outcomes between specific sub-groups. These findings indicate the need to sustain the prevention, identification and management of noncommunicable diseases to reduce the burden of COVID-19.

## Introduction

On 29^th^ December 2019, the WHO reported cases of atypical viral pneumonia in Wuhan, China. The Severe Acute Respiratory Syndrome Coronavirus 2 (SARS-CoV-2) virus was identified as causing this novel coronavirus disease, named COVID-19. The virus is the third zoonotic coronavirus, after SARS-CoV in 2003 and MERS-CoV in 2012 to initiate outbreaks.[1] Genomic studies revealed that horseshoe bats shared 96% of genetic sequences of early SARS-CoV-2 specimens, although it is hypothesised that transmission involved an intermediary host such as a pangolin.[3] Epidemiological links to a live-animal market in Wuhan have been established.[2]

A global pandemic was declared by the World Health Organisation within 74 days. Within 49 weeks, a total of 66.2 million cases and 1.5 million deaths (case fatality rate, 2.3%) were confirmed globally.[4]

Approximately 16% of all cases are estimated to be asymptomatic with 81% presenting with mild clinical characteristics including fever, cough, dyspnoea, and/or fatigue.[5] Categories of disease severity have been outlined by the World Health Organisation, with severe disease more common among older people, those with underlying chronic medical conditions.[6, 7]

Such groups have been classified as ‘vulnerable’ despite limited data on the risk and consequence of infection in Irish cohorts. Due to reliance on non-pharmaceutical interventions (NPIs) to limit transmission, implementing targeted shielding of some vulnerable groups based on risk stratification has been suggested.[8] As the probability of harm may vary over time, based on different NPIs, there is a need to regularly re-assess the relative risk of severe COVID-19 and the impact of infection on outcomes specific groups during different waves of the pandemic.

Using publicly-available data, we aimed to characterise the absolute and the relative risk of adverse clinical outcomes among confirmed COVID-19 cases in the Republic of Ireland. We defined adverse outcomes as hospital or intensive care admission, or death. Specifically, we had two objectives; first, to quantify variation in the frequency of adverse events between two different periods (wave 1 and wave 2), marked by the relaxation of high-level restrictions, and second, to quantify the risk of adverse outcomes associated with (1) age and (2) underlying clinical conditions.

## Methods

We used observational, routinely-collected, publicly-available data available from the Health Protection and Surveillance Centre in the Republic of Ireland. Since the first case of SARS-CoV-2 infection was confirmed on 29^th^ February 2020, data are routinely collected and recorded on the national Computerised Infectious Disease Reporting (CIDR) system, which pre-existed the pandemic and is used for reporting notifiable infectious diseases confirmed in lrish laboratories. Institutional review board approval was not sought for this analysis of publicly available, deidentified data.

Data on age-groups were extracted from reports available up to midnight 14^th^ November 2020, of 67,900 confirmed cases, representing all confirmed cases reported nationally.[9] As the publicly available data stratify cases into specific age-groups, we computed risk metrics associated with these age groups. All cases had a laboratory-confirmed infection (defined as detected on real-time polymerase chain reaction (RT-PCR) testing of oropharyngeal and nasopharyngeal swabs). Data on underlying clinical conditions were extracted from reports available up to midnight 14^th^ November 2020 representing 82.4% (55,946/67,900) of confirmed cases.[10]

We defined two distinct time-points across the observational period. The first, wave 1 (W1) lasted from the date of the first identified SARS-CoV-2 infection to 19 days prior to the date of relaxation of high level restrictions, 81 days in total from 29 Feb 2020 to 20 May 2020). The second, wave 2 (W2) lasted for 177 days from 21 May 2020 to 14 Nov 2020, 17 days before relaxation of the second period of high-level restrictions. Throughout this manuscript, these observational periods will be referred to as wave 1 (W1) and wave 2 (W2), respectively. These dates were chosen to facilitate a comparison of outcomes at similar time-points during the two waves of infections.

Outcomes were classified as hospital admission (i.e. the confirmed case is/was a hospital in-patient on the date of data extraction), intensive care admission (i.e. the confirmed case is/was an in-patient in an intensive care unit on the date of data extraction), death (i.e. the confirmed case was deceased on the date of data extraction).

We calculated the prevalence, and incidence (per 100,000), of laboratory confirmed infections with 2016 census data from the national Central Statistics Office, using the population of each age-group as a denominator (total population = 4,699,608). We also calculated the risk of stated adverse outcomes among confirmed cases with available underlying clinical condition data.

We then calculated the absolute, and relative, risk of stated outcomes among confirmed cases. Relative risk (RR) was calculated by comparing (1) the risk of stated outcomes within each age-group to the risk across all other confirmed cases, and (2) the risk of stated outcomes for cases with specified underlying conditions relative to all other confirmed cases. We also calculated 95% confidence intervals (95% CI) to estimate the precision of these measures.

## Results

The observational period was 258 days (W1: 81 days; W2: 177 days). The number of laboratory confirmed cases was 24,351 and 43,549 in W1 and W2, respectively. Across the population, the overall prevalence of a laboratory confirmed SARS-CoV-2 infections was 1.44% (95% CI 1.43-1.46); prevalence in W1 was 0.52% (95% CI 0.51-0.52) and in W2 was 0.93% (95% CI 0.92-0.93). Healthcare workers accounted for 16.6% (95% CI 16.3-16.9) of confirmed cases (W1 32.0%, 95% CI 31.4-32.6; W2: 8.0%, 95% CI 7.8-8.3).

### Number of Outcomes

A total of 19,198 more infections were reported in W2 than in W1 (43,549 versus 24,351). Despite this, W2 was associated with 1,517 fewer hospital admissions (3,194 versus 1,677), 191 fewer intensive care admissions (393 versus 202), and 935 fewer deaths (1330 versus W2 395).

Among confirmed cases aged ≤64 years old, a total of 21,202 more cases were confirmed in W2 than W1 (39,283 versus 18,081). In W2, those ≤64 years old also had 619 fewer hospital admissions (1,466 versus 827), 152 fewer intensive care admissions (248 versus 96), and 66 fewer deaths (98 versus 32). Similarly, among cases aged ≥65 years old, 1,988 fewer cases were confirmed in W2 (6,246 versus 4,258). In W2 this cohort also experienced 894 fewer hospital admissions (1,745 versus 851), 191 fewer intensive care admissions (393 versus 202), and 935 fewer deaths (1,330 versus 395).

### Incidence of Confirmed Infections

The overall incidence of confirmed infection was 1,426 (95% CI 1416-1437) per 100,000 people. The incidence of confirmed infection was 1.80 times (95% CI 1.77-1.83) higher in W2 (920, 95% CI 911-928 cases per 100,000) than in W1 (512, 95% CI 505-518 cases per 100,000).

In those ≥65 years old, the overall incidence of confirmed infection was 1.19 times (95% CI 1.16-1.21) higher than those ≤64 years old (1653, 95% CI 1621-1685 cases per 100,000 versus 1391, 1380-1403 cases per 100,000). However, this difference was not observed in both observational periods (W1: 2.24, 95% CI 2.18-2.31; W2: 0.71, 95% CI 0.68-0.73).

The incidence of confirmed infection in people aged ≤64 years old was 2.18 times (95% CI 2.14-2.22) higher in W2 than in W1 (957, 95% CI 948-967 versus 439, 95% 432-445 cases per 100,000 respectively). Conversely, among people aged ≥65 years old, the incidence of infection was lower in W2 than in W1 (676, 95% CI 656-696 versus 984, 95% CI 960-1008 cases per 100,000 respectively), resulting in an incidence rate ratio of 0.69 (95% CI 0.66-0.71).

### Risk of Adverse Outcomes Associated with Age

The overall risk of hospitalisation was higher (RR 5.61, 95% CI 5.33-5.91) in cases ≥65 years old (24.7%, 95% CI 23.9, 25.5) compared to those aged ≤65 years old (4.4%, 95% CI 4.2, 4.6),. The risk of ICU admission was also higher RR 3.56 (3.03-4.18) in cases ≥65 years old (2.4%, 95% CI 2.1-2.7) compared to those aged ≤64 years old (0.7%, 95% CI 0.6-0.7). However, the risk of hospital and ICU admission varied considerably between age-groups and observational periods (Table 1&2; Figure 1).

**Table 1.** Risk of Hospital Admission in Confirmed Cases Across Age-Groups (n=67,900) Footnote: Risk metrics presented with 95% confidence intervals. Relative risk (RR) compares absolute risk between age-groups to that of all other cases. Wave 1 (W1) includes 29^th^ February 2020 to 20^th^ May 2020, wave 2 (W2) includes 21^st^ May 2020 to 14^th^ November 2020.

|  | **n** | **Total Risk** | **RR Other Cases** | **W1 Risk** | **W2 Risk** | **W2:W1 RR** |
| --- | --- | --- | --- | --- | --- | --- |
| 0-4 y | 52 | 3.11% (2.28-3.94) | 0.43 (0.33-0.56) | 13.87% (8.08-19.66) | 2.15% (1.42-2.87) | 0.15 (0.09-0.26) |
| 5-14 y | 49 | 1.20% (0.87-1.54) | 0.16 (0.12-0.21) | 6.05% (3.26-8.84) | 0.84% (0.55-1.14) | 0.14 (0.08-0.25) |
| 15-24 y | 151 | 1.28% (1.08-1.48) | 0.15 (0.13-0.18) | 3.97% (3.06-4.88) | 0.81% (0.63-0.98) | 0.20 (0.15-0.28) |
| 24-34 y | 301 | 2.62% (2.33-2.91) | 0.32 (0.29-0.36) | 4.64% (4.00-5.29) | 1.50% (1.22-1.78) | 0.32 (0.26-0.41) |
| 35-44 y | 392 | 3.68% (3.32-4.04) | 0.47 (0.43-0.52) | 5.76% (5.06-6.46) | 2.28% (1.91-2.65) | 0.40 (0.32-0.48) |
| 45-54 y | 621 | 6.01% (5.55-6.47) | 0.81 (0.75-0.88) | 9.79% (8.91-10.67) | 3.23% (2.78-3.68) | 0.33 (0.28-0.39) |
| 55-64 y | 707 | 16.66% (15.54-17.78) | 1.39 (1.29-1.50) | 15.10% (13.85-16.35) | 5.49% (4.81-6.18) | 0.36 (0.31-0.42) |
| 65-74 y | 877 | 22.19% (20.90-23.49) | 3.55 (3.33-3.79) | 32.13% (29.94-34.32) | 14.35% (12.89-15.81) | 0.45 (0.40-0.50) |
| 75-84 y | 1070 | 30.77% (29.24-32.31) | 5.22 (4.92-5.53) | 31.42% (29.52-33.33) | 27.81% (25.34-30.29) | 0.86 (0.88-0.95) |
| ≥85 y | 649 | 21.11% (19.66-22.55) | 3.24 (3.01-3.49) | 20.37% (18.72-22.02) | 23.25% (20.30-26.20) | 1.14 (0.98-1.33) |
| Total | 4871 | 7.17% (6.98-7.37) | - | 13.12% (12.69-13.54) | 3.85% (3.67-4.03) | 0.29 (0.28-0.31) |

**Table 2.** Risk of Intensive Care Admission in Confirmed Cases Across Age-Groups (n=67,900) Footnote: Risk metrics presented with 95% confidence intervals. Relative risk (RR) compares absolute risk between age-groups to that of all other cases. Wave 1 (W1) includes 29^th^ February 2020 to 20^th^ May 2020, wave 2 (W2) includes 21^st^ May 2020 to 14^th^ November 2020.

|  | n | Total Risk | RR Other Cases | W1 Risk | W2 Risk | W2:W1 RR |
| --- | --- | --- | --- | --- | --- | --- |
| 0-4 y | 1 | 0.06% (0.00-0.18) | 0.07 (0.01-0.47) | - | 0.07% (0.00-0.19) | - |
| 5-14 y | 4 | 0.10% (0.00-0.19) | 0.11 (0.04-0.28) | 0.71% (0.00-1.69) | 0.05% (0.00-0.13) | 0.07 (0.01-0.52) |
| 15-24 y | 8 | 0.07% (0.02-0.11) | 0.06 (0.03-0.13) | 0.28% (0.04-0.53) | 0.03% (0.00-0.06) | 0.11 (0.03-0.44) |
| 24-34 y | 17 | 0.15% (0.08-0.22) | 0.14 (0.09-0.23) | 0.34% (0.16-0.52) | 0.04% (0.00-0.09) | 0.12 (0.03-0.41) |
| 35-44 y | 48 | 0.45% (0.32-0.58) | 0.47 (0.35-0.63) | 0.75% (0.49-1.00) | 0.25% (0.13-0.37) | 0.34 (0.19-0.61) |
| 45-54 y | 108 | 1.05% (0.85-1.24) | 1.24 (1.00-1.52) | 1.85% (1.45-2.25) | 0.45% (0.28-0.62) | 0.25 (0.16-0.38) |
| 55-64 y | 158 | 2.14% (1.81-2.47) | 2.96 (2.47-3.55) | 3.63% (2.98-4.29) | 1.04% (0.73-1.34) | 0.29 (0.20-0.40) |
| 65-74 y | 169 | 4.28% (3.65-4.91) | 6.42 (5.39-7.65) | 5.74% (4.65-6.83) | 3.12% (2.40-3.85) | 0.54 (0.40-0.74) |
| 75-84 y | 72 | 2.07% (1.60-2.54) | 2.55 (2.00-3.26) | 1.76% (1.21-2.31) | 2.62% (1.73-3.50) | 1.49 (0.94-2.35) |
| ≥85 y | 8 | 0.26% (0.08-0.44) | 0.29 (0.14-0.58) | 0.26% (0.05-0.47) | 0.25% (0.00-0.61) | 0.97 (0.20-4.79) |
| Total | 595 | 0.88% (0.81-0.95) | - | 1.61% (1.46-1.77) | 0.46% (0.40-0.53) | 0.29 (0.24-0.34) |

**Figure 1.**
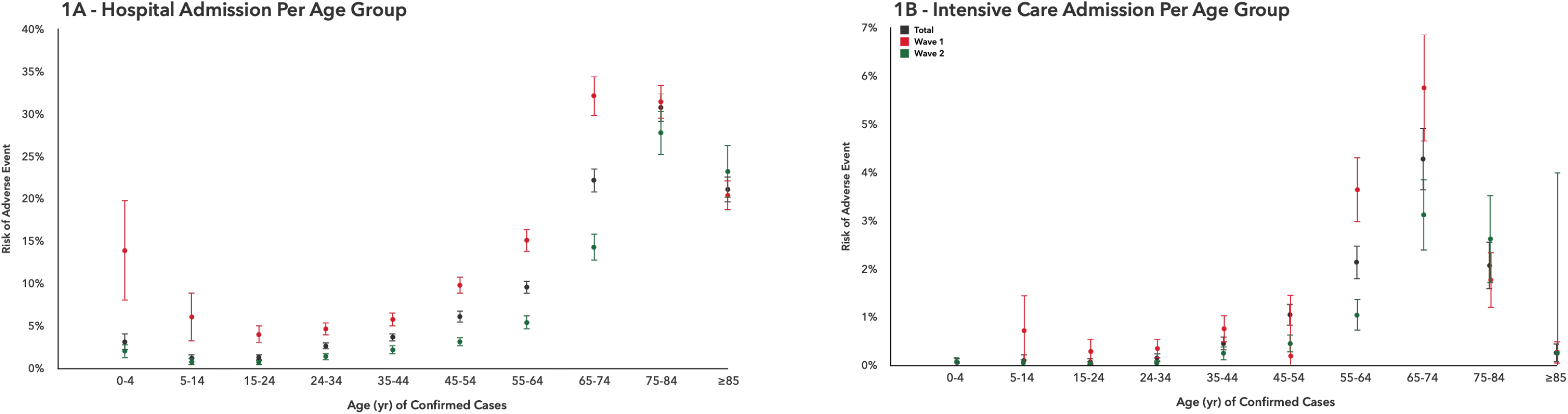
Risk of Hospital and Intensive Care Admission Across Age Groups. Footnote: Figure A reflects hospital admission; Figure B reflects intensive care admissions. Risk metrics presented with 95% confidence intervals. Risk refers to likelihood of event in confirmed cases. Wave 1 (W1) includes 29^th^ February 2020 to 20^th^ May 2020, wave 2 (W2) includes 21^st^ May 2020 to 14^th^ November 2020.

We also found that the overall risk of death was higher RR 60.8 (95% CI 50.9-72.7) in cases ≥65 years old (15.2%, 95% CI14.5-15.9)) compared to those aged ≤64 y (0.25%, 95% CI 0.21-0.29).

### Risk of Adverse Outcomes Associated with Underlying Clinical Conditions

Data on underlying clinical conditions was available for 82% (55,946/67,900) of confirmed cases, 85% (4,120/4,871) of hospital admissions, 100% (593/593) of ICU admissions, and 96% of deaths (1654/1725).

The overall prevalence of ≥1 underlying clinical conditions was 30.5% (95% CI 30.1-30.9) in all confirmed cases. Co-morbidities (≥2 conditions) were recorded for 11.8% (95% CI 11.5-20.0) of cases. Prevalence of underlying conditions was lower (RR 0.40, 95% CI 0.39-0.41) in W2 (20.8%, 95% CI 20.4-21.2) compared to W1 (52.5%, 95% CI 51.8-53.2).

The risk of hospitalisation and ICU admission varied based on underlying clinical conditions across observational periods (Table 3 & 4; Figure 2) with risk of ICU admission remaining relatively stable.

**Table 3.** Risk of Hospital Admission in Confirmed Cases with Underlying Clinical Conditions. Footnote: Risk metrics presented with 95% confidence intervals. Relative risk (RR) compares absolute risk between cases with a specified underlying condition to that of all other cases. Wave 1 (W1) includes 29^th^ February 2020 to 20^th^ May 2020, wave 2 (W2) includes 21^st^ May 2020 to 14^th^ November 2020.

|  | n | Total Risk | RR Other Cases | OP1 Risk | OP2 Risk | OP2:OP1 RR |
| --- | --- | --- | --- | --- | --- | --- |
| Chronic heart disease | 1214 | 34.66% (33.08-36.23) | 2.57 (2.42-2.74) | 37.18% (35.26-39.09) | 28.83% (26.10-31.56) | 0.78 (0.70-0.86) |
| Chronic respiratory disease | 776 | 18.96% (17.76-20.16) | 1.09 (1.01-1.17) | 27.19% (25.15-29.23) | 12.32% (10.97-13.68) | 0.45 (0.40-0.52) |
| Diabetes mellitus | 618 | 32.25% (30.16-34.35) | 1.96 (1.82-2.12) | 38.81% (35.90-41.72) | 22.62% (19.87-25.38) | 0.58 (0.51-0.67) |
| Cancer/malignancy | 456 | 39.86% (37.02-42.70) | 2.46 (2.27-2.66) | 48.58% (44.68-52.47) | 29.10% (25.17-33.04) | 0.60 (0.51-0.70) |
| Chronic neurological disease | 440 | 27.28% (25.10-29.45) | 1.62 (1.49-1.77) | 30.56% (27.71-33.40) | 21.82% (18.53-25.11) | 0.71 (0.60-0.85) |
| Chronic kidney disease | 344 | 43.05% (39.62-46.49) | 2.60 (2.38-2.83) | 49.89% (45.38-54.41) | 33.23% (28.13-38.33) | 0.67 (0.56-0.80) |
| Asthma requiring medication | 188 | 32.92% (29.07-36.78) | 1.90 (1.69-2.15) | 33.95% (29.48-38.43) | 29.79% (22.24-37.34) | 0.88 (0.66-1.17) |
| BMI ≥ 40 | 161 | 47.92% (42.57-53.26) | 4.27 (3.79-4.81) | 43.03% (36.82-49.25) | 60.87% (50.90-70.84) | 1.41 (1.14-1.76) |
| Immunodeficiency including HIV | 146 | 23.14% (19.85-26.43) | 1.31 (1.13-1.52) | 28.13% (23.43-32.82) | 16.85% (12.45-21.24) | 0.60 (0.44-0.82) |
| Chronic Liver Disease | 86 | 35.10% (29.13-41.08) | 2.00 (1.68-2.38) | 40.69% (32.69-48.69) | 27.00% (18.30-35.70) | 0.66 (0.45-0.97) |

**Table 4.** Risk of Intensive Care Admission in Confirmed Cases with Underlying Clinical Conditions. Footnote: Risk metrics presented with 95% confidence intervals. Relative risk (RR) compares absolute risk between cases with a specified underlying condition to that of all other cases. Wave 1 (W1) includes 29^th^ February 2020 to 20^th^ May 2020, wave 2 (W2) includes 21^st^ May 2020 to 14^th^ November 2020.

|  | n | Total Risk | RR Other Cases | W1 Risk | W2 Risk | W2:W1 RR |
| --- | --- | --- | --- | --- | --- | --- |
| Chronic heart disease | 277 | 7.91% (7.01-8.80) | 4.34 (3.67-5.13) | 7.98% (6.90-9.05) | 7.75% (6.14-9.36) | 0.97 (0.76-1.24) |
| Chronic respiratory disease | 156 | 3.81% (3.23-4.40) | 1.34 (1.12-1.61) | 5.36% (4.33-6.39) | 2.56% (1.91-3.21) | 0.48 (0.35-0.66) |
| Diabetes mellitus | 162 | 8.26% (7.04-9.48) | 3.44 (2.88-4.12) | 8.73% (7.04-10.41) | 7.69% (5.94-9.45) | 0.88 (0.65-1.19) |
| Cancer/malignancy | 75 | 6.56% (5.12-7.99) | 2.32 (1.83-2.94) | 6.80% (4.84-8.77) | 6.25% (4.15-8.35) | 0.92 (0.59-1.43) |
| Chronic neurological disease | 28 | 1.74% (1.10-2.37) | 0.54 (0.37-0.79) | 1.29% (0.59-1.99) | 2.48% (1.24-3.72) | 1.92 (0.92-4.01) |
| Chronic kidney disease | 59 | 7.38% (5.57-9.20) | 2.58 (1.99-3.35) | 6.16% (3.99-8.33) | 9.15% (6.03-12.27) | 1.49 (0.91-2.43) |
| Asthma requiring medication | 74 | 12.96% (10.20-15.71) | 4.75 (3.77-5.98) | 10.70% (7.78-13.62) | 19.86% (13.27-26.44) | 1.86 (1.21-2.85) |
| BMI ≥40 | 107 | 31.85% (26.86-36.83) | 12.77 (10.63-15.33) | 26.64% (21.09-32.19) | 45.65% (35.47-55.83) | 1.71 (1.26-2.33) |
| Immunodeficiency including HIV | 36 | 5.71% (3.90-7.51) | 1.92 (1.38-2.67) | 6.82% (4.18-9.45) | 4.30% (1.92-6.68) | 0.63 (0.32-1.24) |
| Chronic Liver Disease | 19 | 7.76% (4.41-11.10) | 2.58 (1.66-4.01) | 8.28% (3.79-12.76) | 7.00% (2.00-12.00) | 0.85 (0.35-2.07) |

**Figure 2.**
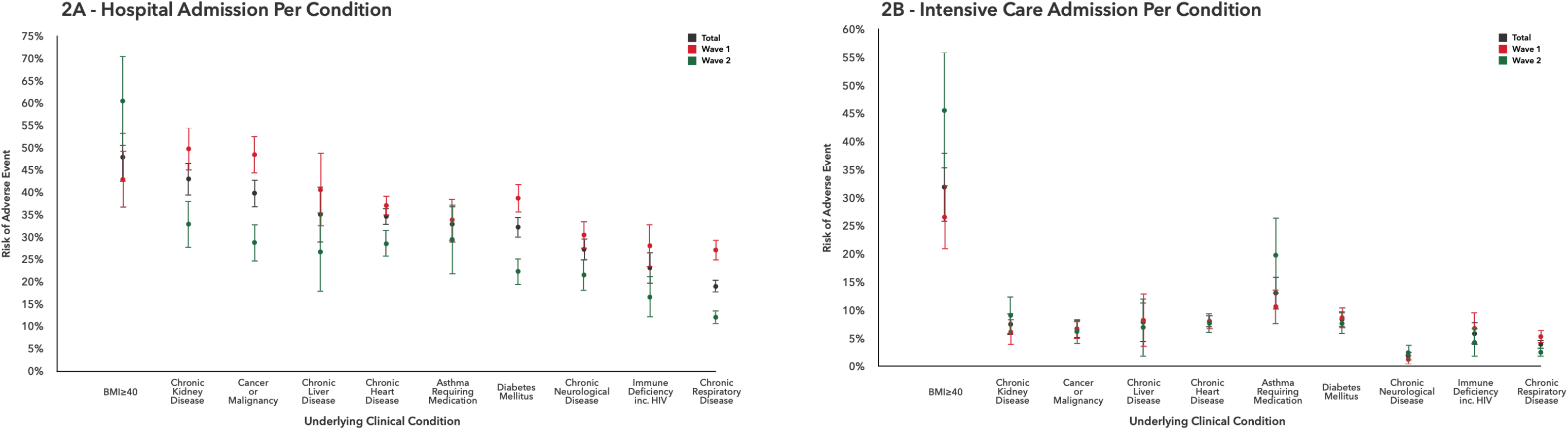
Risk of Hospital and Intensive Care Admission with Underlying Clinical Conditions. Footnote: Figure A reflects hospital admission; Figure B reflects intensive care admissions. Risk metrics presented with 95% confidence intervals. Risk refers to likelihood of event in confirmed cases. Wave 1 (W1) includes 29^th^ February 2020 to 20^th^ May 2020, wave 2 (W2) includes 21^st^ May 2020 to 14^th^ November 2020.

## Discussion

Despite frequent references in the media and scientific literature to more adverse consequences for ‘vulnerable’ or ‘high-risk’ individuals, to our knowledge this is the first attempt to quantify the risk of adverse clinical outcomes in Irish populations with COVID-19.

Compared to W1, W2 was associated with (1) increased incidence of confirmed infections in people ≤65 years and (2) reduced prevalence of underlying conditions among confirmed cases. These findings indicate the need for (1) bespoke mitigation strategies reflecting the consequences of infection for specific age cohorts on hospital services, and (2) ongoing prevention, identification and management of diseases associated with increased risk of adverse outcomes following COVID-19 infections.

The risk of adverse outcomes is significantly higher among people with underlying conditions, remaining relatively constant across waves of infections for those with a BMI ≥40 or asthma requiring medication. Also, the risk of ICU admission did not reduce in W2 among cases with underlying conditions, with the exception of chronic respiratory disease. Despite comorbidities being reported for only 12% of people with COVID-19 in the Republic of Ireland, we suggest a greater focus on risk management, including undisrupted access to healthcare, for people with multiple diseases.

As these conditions are not age-dependent (e.g. BMI ≥40, diabetes mellitus, asthma, cancer) it is important to address the biopsychosocial aspects driving the risk and burden of these diseases. As modifiable factors play a large role in the pathogenesis of most diseases associated with adverse outcomes, the disease burden is likely amplified when SARS-CoV-2 spreads among societies that have not persistently promoted all aspects of population health. This requires that responses to COVID-19 embed promotion of healthy lifestyles and early detection and management of non-communicable disease (NCD) alongside infection control measures. Although prevention of NCD may not be an immediate priority, failure to deal with the drivers of these diseases inevitably increases the harm that novel pathogens will have in the future.

Additionally, early detection of suspected SARS-CoV-2 infection, particularly in those at increased risk of adverse outcomes, should be prioritised. In localised regions or facilities, this could aid infection prevention and control measures while simultaneously enhancing patient management, resource utilisation and early-targeted vaccination implementation. To achieve this, easy-to-use advanced surveillance and triage systems are required, particularly where immediate access to testing is limited.[11]

Our analysis does have limitations. As we did not have access to individual level data we were unable to fully adjust analyses for additional potential confounders and, in particular, to account for interactions between age and underlying comorbidities. In addition, as many comorbidities occur in clusters, we were unable to model which underlying condition(s) most contributed to worse outcomes.

Given the high proportion of confirmed infections acquired by healthcare-worker (17%) data on the proportion of cases that were inpatients or close contacts of healthcare-workers would help to more accurately gauge the rate of transmission and risk of adverse outcomes in those acquiring infection in healthcare facilities.

Individual level, de-identified data would assist in identifying innovative responses to support targeted strategies to reduce the risk of transmission of SARS-CoV-2 infection in those with co-morbidities, thereby reducing the burden of COVID-19 on the local healthcare systems.

## Data Availability

These data were derived from the following resources available in the public domain: https://www.hpsc.ie. The data that support the findings of this study are available from the corresponding author, MR.

https://www.hpsc.ie/a-z/respiratory/coronavirus/novelcoronavirus/casesinireland/epidemiologyofcovid-19inireland/may2020/COVID-19%20NPHET%20report%2020200522_%20Website.pdf

https://www.hpsc.ie/a-z/respiratory/coronavirus/novelcoronavirus/casesinireland/epidemiologyofcovid-19inireland/november2020/COVID-19_Daily_epidemiology_report_(NPHET)_20201116%20-%20website.pdf

https://www.hpsc.ie/a-z/respiratory/coronavirus/novelcoronavirus/surveillance/underlyingconditionsreports/

